# Multi-Modal Wearable and Biological Data Integration Reveals Physiological Signatures of Weight Variation in Overweight and Obese Adults: An AI4Food Machine Learning Study

**DOI:** 10.64898/2026.11.03.26367178

**Authors:** Hicham Attioui, Blanca Lacruz-Pleguezuelos, Enrique Carrillo de Santa Pau

**Author notes:** Corresponding authors: Hicham Attioui,; Enrique Carrillo de Santa Pau.

## Abstract

This study is part of the AI4Food project, which aims to identify the physiological and behavioral factors influencing weight variation in overweight and obese individuals through integrative analysis of multi-source data. A 30-day crossover intervention was conducted on 93 participants at IMDEA Nutrition Institute, combining a traditional phase (questionnaires, food diaries) and a digital phase (Fitbit Sense Smartwatch, FreeStyle Libre 2 glucose sensor). The collected data encompassed sleep, glucose, skin temperature, nutrition, and gut microbiome information. After rigorous preprocessing and alignment by intervention days, unsupervised clustering, association analysis, and machine learning models were applied to uncover physiological patterns linked to weight dynamics. Results showed greater weight reduction during the digital phase in Group 2, while between-group differences were not significant. Significant associations were observed between nocturnal respiratory rate, skin temperature, and weight variation. Under participant-level five-fold cross-validation, Random Forest models achieved mean accuracies of 64.3% for thermoregulation and 61.7% for glucose phenotype prediction, with RMSSD, restlessness, and nocturnal respiratory rate among the most influential predictors. In contrast, microbiome composition remained almost stable over the short intervention period. These findings support the potential of AI-driven integration of wearable and nutritional data to stratify metabolic phenotypes and advance personalized nutrition strategies in obesity management.

## INTRODUCTION

Obesity is one of the most pressing global health challenges of the twenty-first century. According to recent projections from the World Obesity Atlas 2023 (World Obesity Federation, 2023), nearly 25% of the world’s adult population is expected to live with obesity by 2035, highlighting the accelerating nature of this epidemic. This condition is associated with an increased risk of metabolic, cardiovascular, and inflammatory diseases and represents a major burden for healthcare systems worldwide. Traditionally regarded as the consequence of an imbalance between caloric intake and energy expenditure, obesity is now understood as a multifactorial disorder influenced by genetic, hormonal, behavioral, and environmental determinants (Blüher, 2019). Among these, sleep has emerged as a key physiological regulator of metabolic balance. Numerous studies have demonstrated that short or poor-quality sleep alters appetite-related hormones, disrupts glucose metabolism, and promotes excessive food intake, ultimately favoring weight gain and metabolic dysregulation (Reutrakul & Van Cauter, 2018).

Over the last decade, technological advances have profoundly transformed the investigation of these mechanisms. The growing use of wearable sensors allows for the continuous and non-invasive monitoring of physiological parameters such as heart rate variability (HRV), oxygen saturation (SpO2), respiratory rate, skin temperature, and sleep stages in real-world conditions (Dunn et al., 2018). These digital devices generate high-frequency longitudinal data that complement traditional clinical and nutritional assessments, enabling a more comprehensive characterization of metabolic health. When integrated with dietary, biological, and behavioral information, such datasets form the foundation for precision nutrition, where interventions can be adapted to each individual’s physiological and behavioral profile (Zeevi et al., 2015).

Within this framework, the AI4Food project was established to integrate wearable, nutritional, and clinical data to improve understanding of weight regulation and individual metabolic responses. Using a crossover design that alternated traditional and digital monitoring phases within the same participants, the study allowed for a direct comparison between the two data-collection modalities while minimizing interindividual variability. The clinical feasibility and primary weight-loss outcomes of this same crossover intervention (n = 93), including changes in weight, BMI, visceral fat, cholesterol, and HbA1c, have been reported separately (Lacruz-Pleguezuelos et al., 2025). The present study builds on that work by focusing on the underlying physiological and behavioral phenotypes captured through wearable-derived sleep, thermoregulation, and glucose data.

The integration of such complex and heterogeneous data requires advanced computational tools. Machine learning and multivariate modeling enable the identification of hidden structures and relationships between physiological variables, while clustering techniques reveal distinct metabolic or behavioral phenotypes (Metwally et al., 2026). Predictive modeling approaches can assess whether sleep-derived physiological parameters contain information associated with metabolic phenotypes such as glucose regulation or thermoregulation. Moreover, recent developments in explainable artificial intelligence (XAI), such as SHapley Additive exPlanations (SHAP), provide interpretable frameworks that clarify the contribution of each physiological feature to model predictions, thereby facilitating biological interpretation (Lundberg & Lee, 2017).

The present study aims to identify physiological signatures associated with weight variation through the integration of wearable-derived, nutritional, biological, and gut microbiome data collected during the AI4Food intervention. By examining the relationships among sleep quality, thermoregulation, glucose variability, nutritional behavior, and gut microbiome composition, this work seeks to advance understanding of how digital biomarkers reflect metabolic flexibility and responsiveness to dietary interventions. Through the combined use of data-driven modeling and biological interpretation, the study contributes to the development of AI-assisted precision nutrition strategies aimed at improving weight management and metabolic health in individuals with obesity.

## MATERIALS & METHODS

### Study Design and Context

This work was conducted within the framework of the AI4Food project, a clinical– digital study designed to explore the determinants of weight variation through the integration of multi-source data. The study followed a 30-day crossover design involving 93 participants with overweight or obesity, recruited according to specific inclusion criteria (age ≥18 years, BMI between 27 and 35 kg/m², ability to comply with the protocol, and willingness to use wearable devices). The clinical feasibility and primary outcomes of this intervention, including changes in weight, BMI, visceral fat, cholesterol, and HbA1c, have been reported separately (Lacruz-Pleguezuelos et al., 2025). Participants were randomly assigned to two groups: Group 1 began with a manual phase followed by a digital phase, while Group 2 followed the reverse order. Each phase lasted 15 days, allowing all participants to experience both traditional and technology-assisted interventions. This structure enabled comparison between data collection modalities while minimizing interindividual variability.

The intervention combined nutritional guidance, self-monitoring, and digital tracking. During the manual phase, participants recorded food intake and completed validated questionnaires. During the digital phase, they were equipped with wearable sensors providing continuous physiological measurements. The study was conducted at the IMDEA Nutrition Institute (Madrid, Spain) and was approved by the Research Ethics Committee of the IMDEA Nutrition Foundation (protocol IMD PI-052, approved April 5, 2022). The trial was registered at ClinicalTrials.gov (NCT05807243). All participants provided informed consent before enrollment.

**Figure 1.**
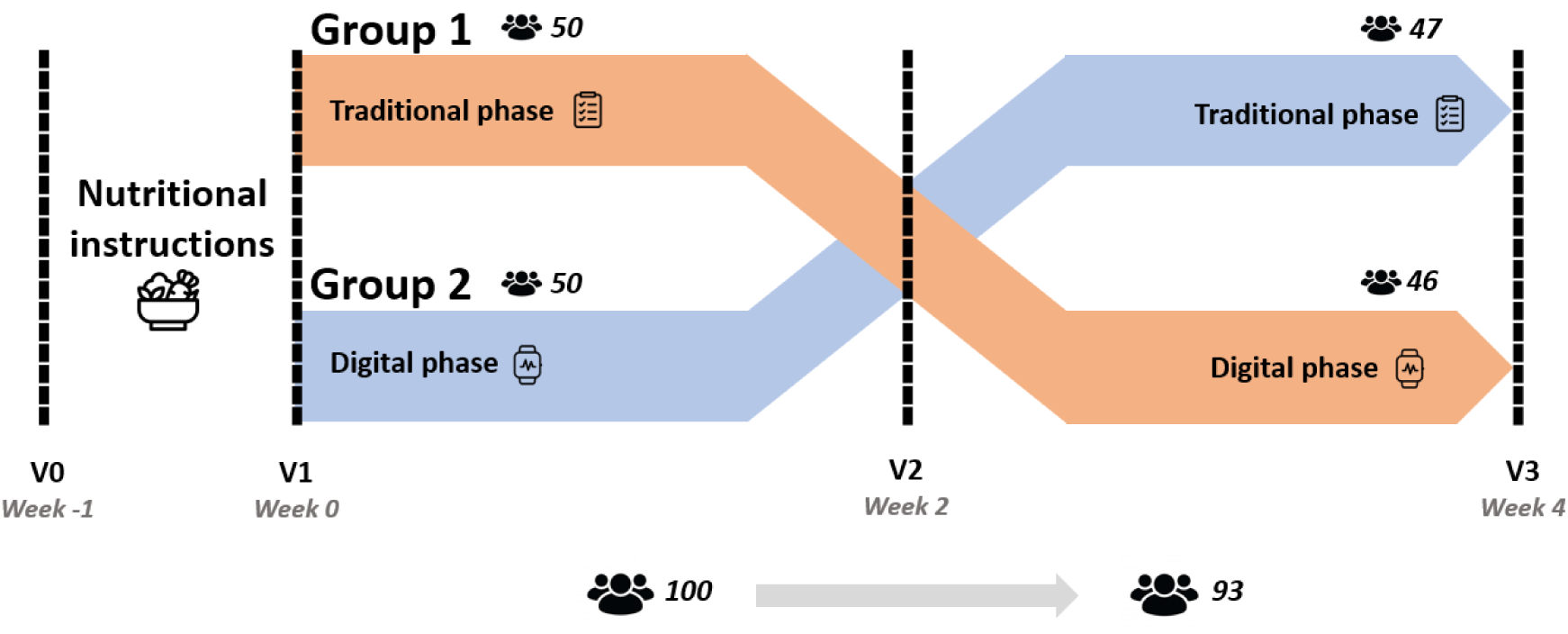
An overview of the AI4Food crossover design. The 30-day intervention followed a crossover structure in which participants alternated between traditional data collection (manual questionnaires and food diaries) and digital monitoring using wearable sensors. Group 1 began with the traditional phase, while Group 2 started with the digital phase (Romero-Tapiador et al., 2023).

### Data Collection

The study integrated **digital**, **traditional**, and **clinical** datasets into a unified structure (AI4FoodDB, Romero-Tapiador et al., 2023).

### Digital data

Included continuous physiological signals from two wearable devices:

- Fitbit Sense Smartwatch (Fitbit LLC): The Fitbit Sense is a commercial wearable device that runs the Fitbit OS operating system and connects to a mobile phone via the Fitbit app (compatible with iOS 12.2 or Android 8.0 and higher). The device can be worn on either wrist, weighs approximately 46 g, and has a battery life of 4-6 days. It incorporates a series of sensors for continuous health monitoring: an optical heart rate sensor, an electrodermal activity (EDA) sensor, inertial sensors, a skin temperature sensor, a 1-lead electrocardiogram (ECG), and sleep quality detection algorithms. The watch automatically estimated sleep stages (light, deep, REM), total sleep duration, respiratory rate, heart rate variability (HRV, RMSSD), and nocturnal skin temperature variations (Romero-Tapiador et al., 2023).
- FreeStyle Libre 2 Glucose Sensor (Abbott Laboratories): The FreeStyle Libre 2 is a continuous glucose monitoring (CGM) device that measures interstitial glucose every 15 minutes. The sensor measures 35×5 mm, weighs approximately 5 g, and must be replaced every 14 days. It is placed on the back of the upper arm and transmits data to a smartphone via Near Field Communication (NFC), returning up to 8 hours of collected data per scan. This device provides a minimally invasive, high-frequency measure of interstitial glucose, enabling identification of daily glycemic patterns and their association with sleep and nutritional behaviors (Romero-Tapiador et al., 2023; Hoss & Budiman, 2017).

### Traditional data

Comprised sleep questionnaires, food diaries, and lifestyle surveys collected during the non-digital phase. The Oviedo Sleep Questionnaire (OSQ) was used to assess subjective sleep satisfaction, insomnia, and hypersomnia (Bobes et al., 2000). Dietary intake was assessed using three-day dietary records, in which participants recorded all food and beverages consumed over three consecutive days (including one weekend day). Records were completed by hand and subsequently entered into the DIAL software (v3.15) to estimate total energy intake and macro-and micronutrient composition (Ortega et al., 2021).

### Clinical and biological data

Obtained at three main visits (baseline V1, mid-intervention V2, post-intervention V3) and included anthropometric measures (height, waist-to-hip ratio, BMI), metabolic biomarkers (glucose, insulin, lipids, CRP, adiponectin), and gut microbiome profiles obtained by shotgun metagenomic sequencing of stool samples collected at V1 and V3. Body weight was additionally recorded at a pre-baseline visit (V0) to establish each participant’s pre-intervention weight, and again at V2 and V3 to track weight change across intervention phases.

All data were anonymized and harmonized within the AI4Food database to facilitate multi-modal integration (Romero-Tapiador et al., 2023).

### Data Preprocessing

A standardized data-cleaning and harmonization pipeline was implemented using Python 3.11 and GraphPad Prism 8.4. Data processing and integration were performed using pandas, NumPy, and SciPy, while visualization and quality control were conducted using matplotlib and seaborn.

### Wearable data (Fitbit Sense)

Daily aggregates were exported from the Fitbit API and aligned to each participant’s intervention calendar to account for differences in starting dates. Only measurements falling within each participant’s digital-monitoring phase were retained. Variables with substantial missingness were excluded; remaining missing values were examined against physiological plausibility thresholds. Outliers were identified using the interquartile range criterion and verified manually against physiologically implausible values. Following integration, repeated participant-day records were consolidated to yield a single observation per participant per day, resulting in 261 participant-day observations from 55 participants for the thermoregulation dataset.

#### Continuous glucose monitoring data (FreeStyle Libre 2)

Interstitial glucose readings (one per 15 minutes) were aligned to intervention days and filtered to retain only measurements from the digital-monitoring phase. Missing scans and outlier glucose values were examined against physiological reference ranges. After consolidation of repeated records, the glucose prediction dataset comprised 144 participant-day observations from 44 participants.

### Gut microbiome data

Stool samples were collected at visits V1 and V3 (97 samples at V1, 91 at V3) to characterize gut microbiome composition and functionality via shotgun metagenomics and untargeted metabolomics. Sample collection and sequencing procedures followed established protocols, and taxonomic profiles were computed at the genus and phylum level and matched to participant clinical and wearable data by visit timepoint (Romero-Tapiador et al., 2023).

### Clinical and anthropometric data

Biomarkers and anthropometric measures collected at visits V0-V3 were verified for physiological plausibility. Variables with substantial missingness across participants were excluded from downstream analyses.

### Dietary data

Three-day dietary records were manually entered into DIAL software (Ortega et al., 2021). Nutrient variables were checked for completeness and implausible values were flagged and reviewed.

### Statistical and Computational Analyses

Descriptive statistics were computed for all variables. Within-subject comparisons across intervention phases (e.g., OSQ and weight change between visits in the same participants) were assessed using paired t-tests or Wilcoxon signed-rank tests, depending on distribution normality (Shapiro-Wilk test). Between-group comparisons were assessed using independent t-tests or Wilcoxon rank-sum tests. Categorical variables were compared using Chi-square tests. Given the number of statistical comparisons performed across physiological domains, p-values were not adjusted for multiple comparisons; results should therefore be interpreted as hypothesis-generating rather than confirmatory. To explore global variability in dietary patterns, Principal Component Analysis (PCA) was applied to normalized nutritional variables.

### Clustering

Unsupervised clustering was used to characterize physiological, nutritional, and weight-response patterns. K-means clustering was applied separately to the relevant physiological or nutritional datasets, and the number of clusters was selected using the elbow method together with inspection of cluster separation and interpretability. Cluster assignments were visualized longitudinally using participant- by-day heatmaps to evaluate the stability and temporal variation of physiological phenotypes.

### Inter-cluster association

To investigate relationships between physiological domains, contingency matrices were built comparing cluster memberships across modalities (e.g., sleep vs. glucose, temperature vs. weight clusters). Statistical significance was tested using the Chi-square test and, when appropriate, Spearman correlations.

### Predictive modeling

Two supervised classification analyses were conducted to predict thermoregulation and glucose phenotypes using Random Forest (Breiman, 2001) and multilayer perceptron (MLP) models (Schmidhuber, 2015). Fifteen raw physiological variables were used for thermoregulation prediction, excluding direct and derived temperature variables to avoid target-related information leakage. For glucose prediction, the same variables were used together with nightly temperature and baseline-relative temperature variability, while thermoregulation cluster membership was excluded. Model performance was evaluated using five-fold participant-level cross-validation with StratifiedGroupKFold implemented in scikit-learn (Pedregosa et al., 2011), ensuring that observations from the same participant were never present in both training and validation sets. Precomputed normalized variables were not used as predictors; instead, raw physiological variables were retained so that preprocessing could be performed independently within each cross-validation training fold. Random Forest models were trained directly on raw predictor values, while MLP inputs were standardized within each training fold only and applied to the held-out observations. Performance was assessed using accuracy, precision, recall, F1-score, and ROC-AUC and summarized as mean ± standard deviation across folds. Precision, recall, and F1-score were calculated using the high-temperature or high-glucose phenotype as the positive class. SHAP values were calculated from out-of-fold Random Forest predictions to identify the most influential features.

Although the complete intervention cohort comprised 93 participants, the effective sample size differed across predictive analyses because only participants with matched, quality-controlled physiological measurements could be included.

## RESULTS

### Weight Variation Across Phases

The intervention resulted in a significant overall reduction in body weight, with participants achieving a mean reduction of 2.00 kg over the full 30-day program (V1: 84.99 ± 13.69 kg; V3: 82.72 ± 13.32 kg; p < 0.001), confirming the success of the weight loss intervention (Lacruz-Pleguezuelos et al., 2025).

Weight was measured at three clinical visits (V0, V2, V3) to assess the impact of the two monitoring modalities. In Group 2, participants lost significantly more weight during the digital phase (V0-V2: −1.51 ± 1.08 kg) than during the traditional phase (V2-V3: −0.79 ± 1.11 kg; p < 0.01). Group 1 showed a similar directional pattern (traditional phase V0-V2: −1.03 ± 1.49 kg; digital phase V2-V3: −1.24 ± 1.22 kg), but this within-group difference did not reach significance (p = 0.42). Between-group comparisons at each phase were also not significant (V0-V2: p= 0.08; V2-V3: p= 0.07).

**Figure 2.**
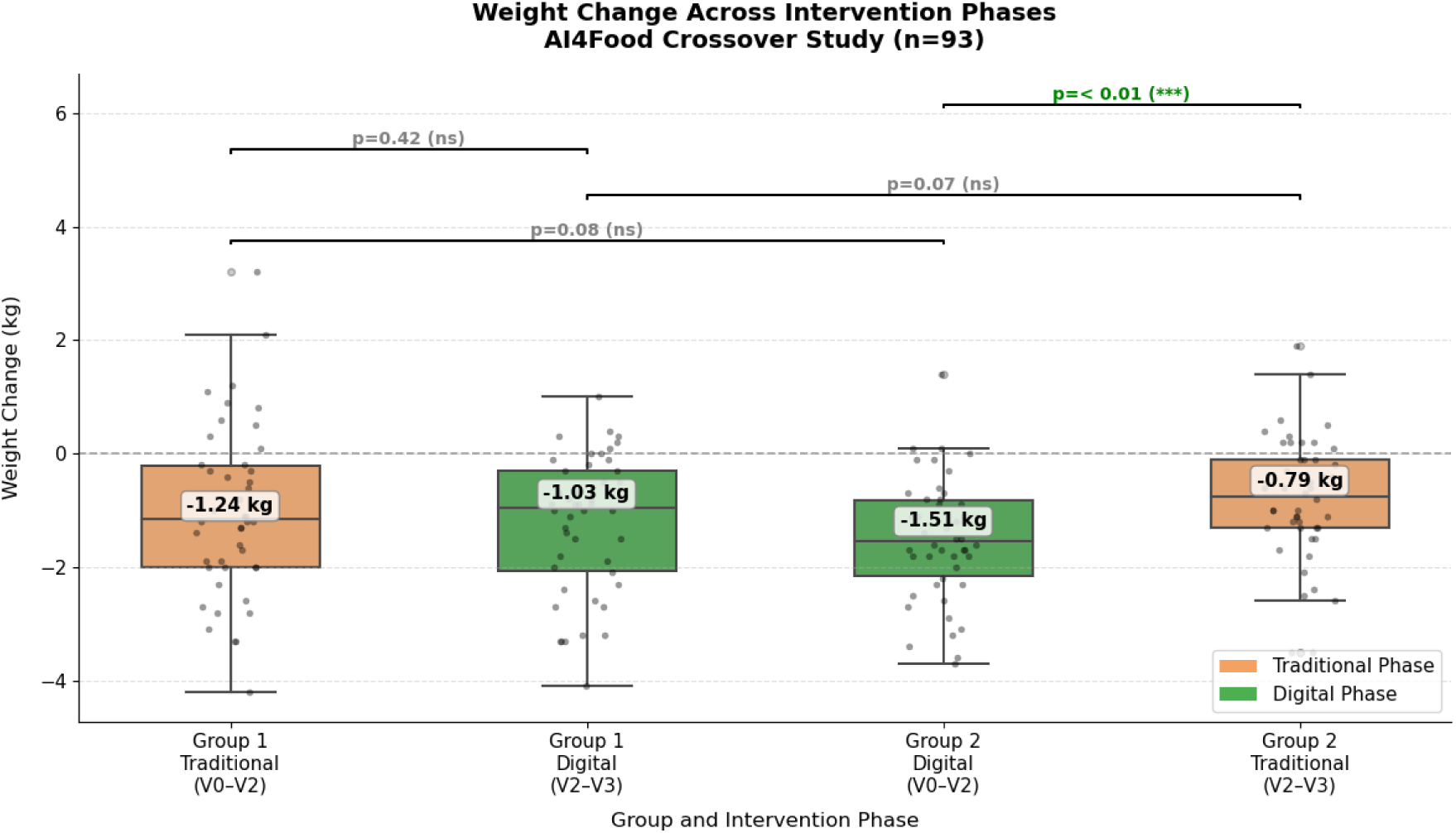
Comparison of weight change across intervention phases. Boxplots show the distribution of individual weight changes (kg) over each phase, colored by intervention type: Traditional (orange) and Digital (green). Mean weight change values are displayed inside each box. Group 1 began with the Traditional phase (V0-V2) followed by the Digital phase (V2-V3), while Group 2 followed the reverse order.

### Weight-Based Clustering

To capture heterogeneous weight-loss responses, participants with complete weight data at both V0 and V3 (n = 92) were grouped into three weight-variation clusters based on total change over this period.

**Figure 3.**
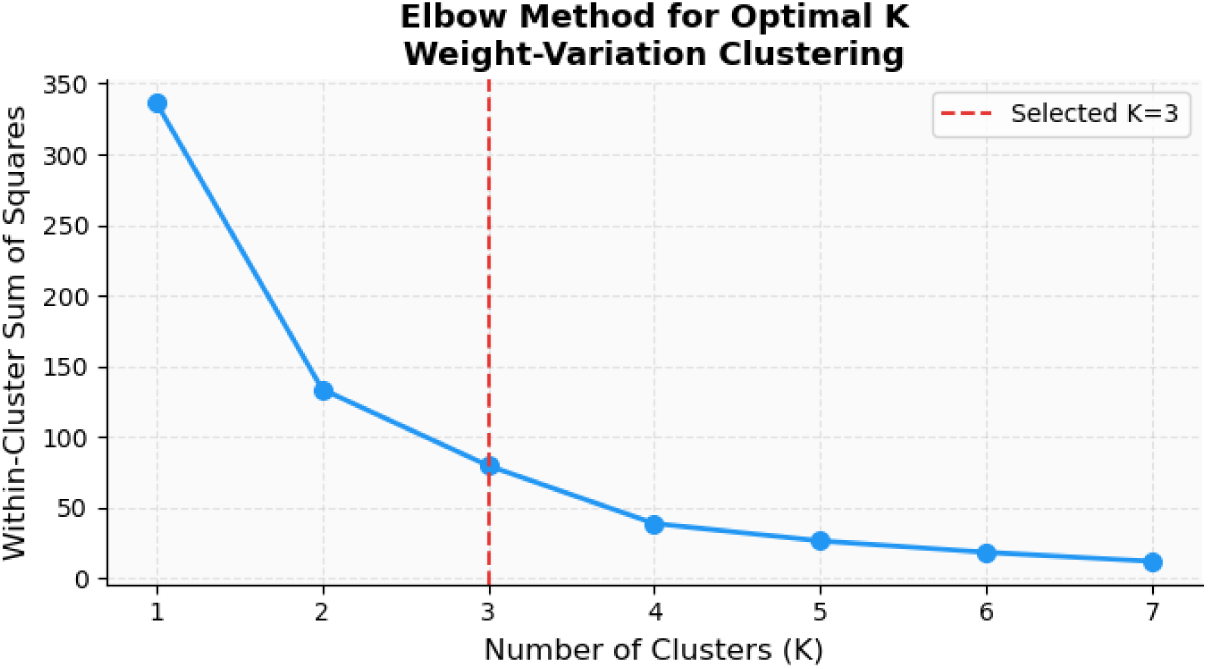
Elbow method for optimal cluster selection in weight-variation clustering.

The optimal number of clusters (K = 3) was confirmed by the elbow method, balancing granularity and interpretability:

**W_Cluster 0** (n = 24): Marked responders (−4.68 ± 1.18 kg)

**W_Cluster 1** (n = 58): Moderate responders (−1.86 ± 0.80 kg)

**W_Cluster 2** (n = 10): Non-responders or weight gain (+1.01 ± 1.13 kg)

The resulting clusters represented marked responders, moderate responders, and participants with minimal weight loss or weight gain.

**Figure 4.**
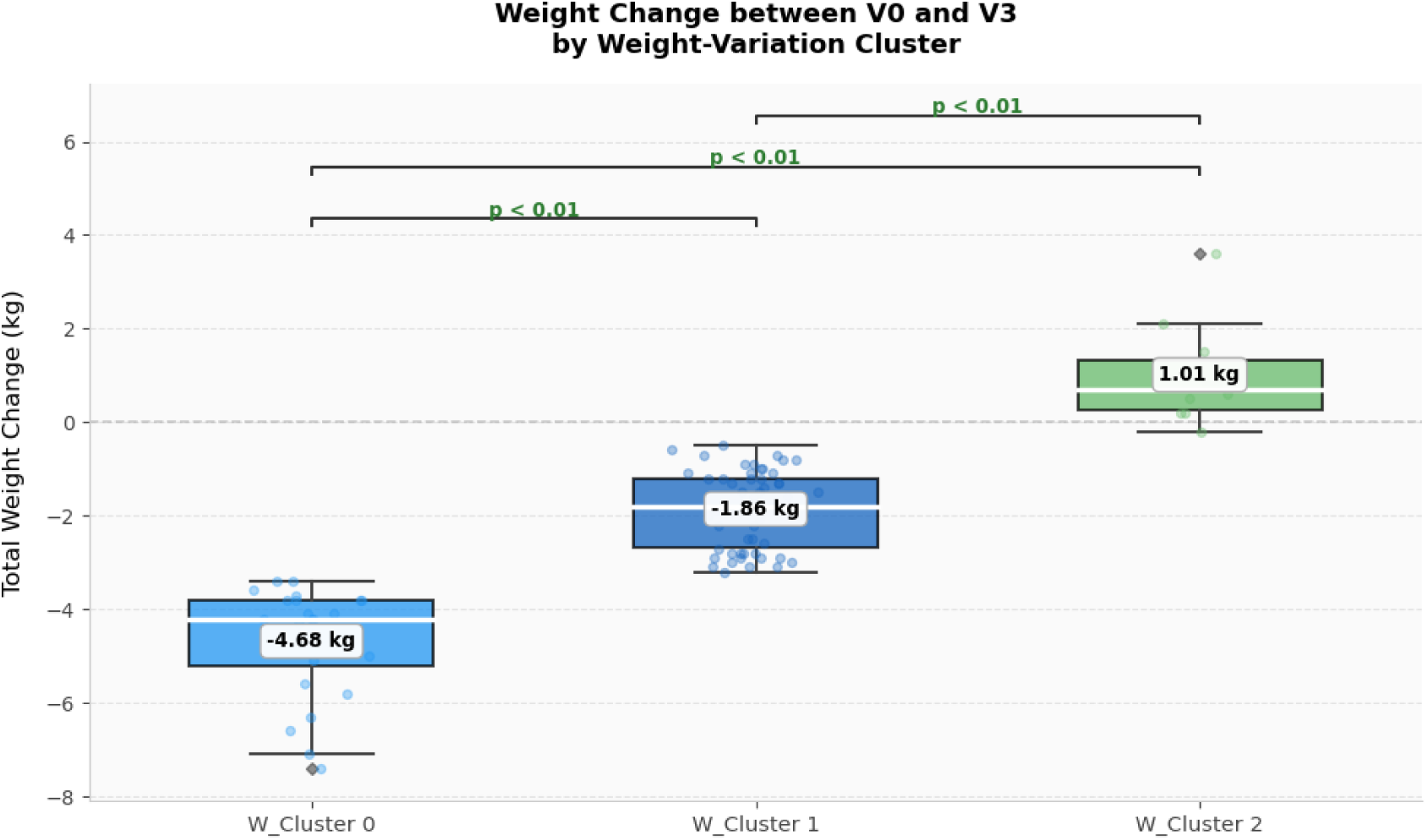
Total weight change between baseline (V0) and post-intervention (V3) across clusters. Boxplots show distinct weight-response groups: W_Cluster 0 (−4.68 kg), W_Cluster 1 (−1.86 kg), and W_Cluster 2 (+1.01 kg).

### Sleep Questionnaire (OSQ)

The OSQ provides a global index of subjective sleep quality, where higher scores indicate greater sleep complaints. In Group 1, OSQ scores showed a slight decrease between V1 and V2 (27.1 ± 6.5 to 26.6 ± 6.9; p = 0.45), suggesting a modest but non-significant improvement in perceived sleep quality during the traditional phase. In Group 2, a significant decrease in OSQ scores was observed between V2 and V3 (27.7 ± 5.8 to 26.0 ± 6.0; p = 0.03), indicating a meaningful improvement in subjective sleep quality following the traditional phase. Overall, both groups showed a trend toward better perceived sleep quality across the intervention, with the effect reaching statistical significance only in Group 2.

**Figure 5.**
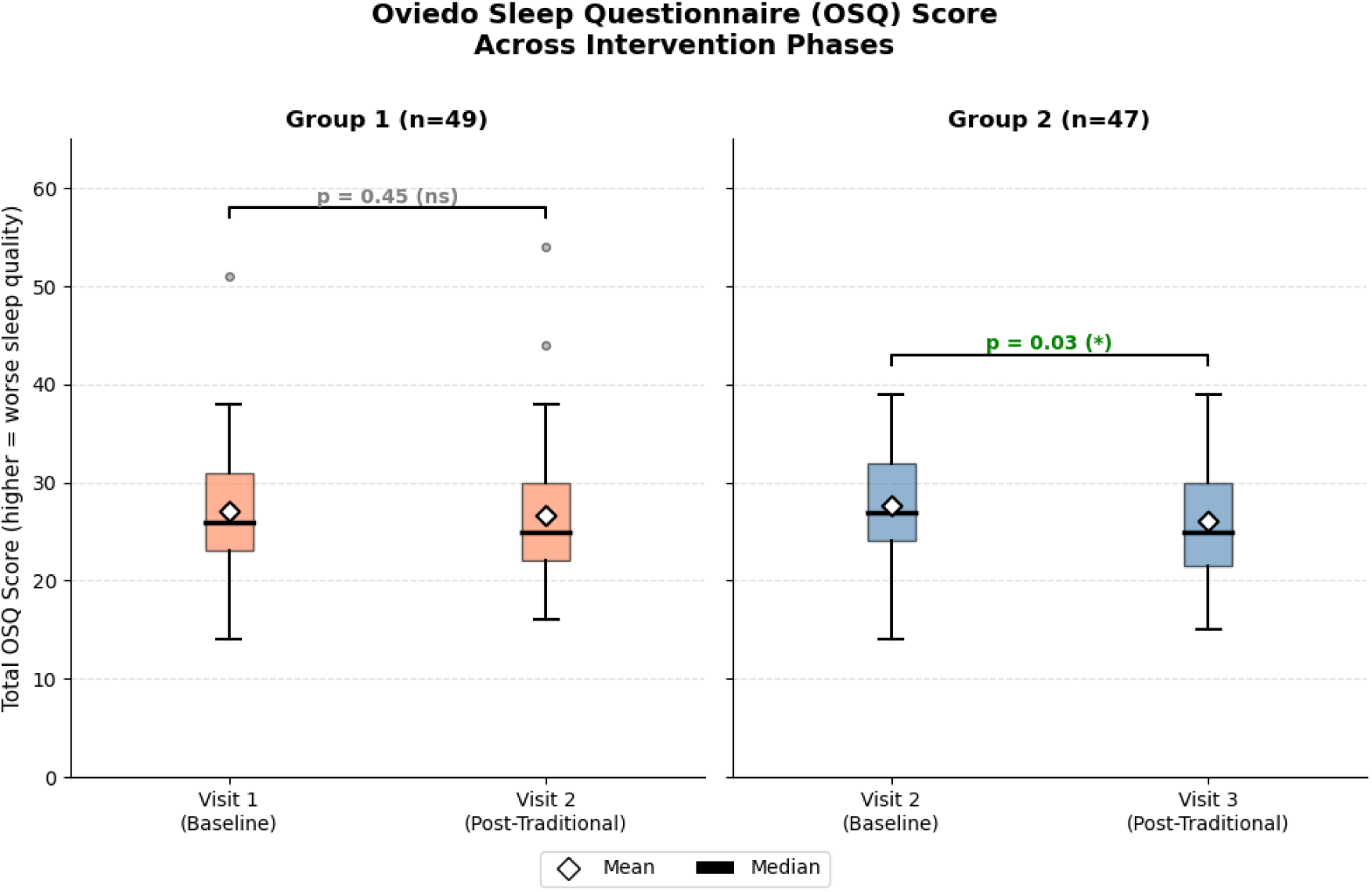
Evolution of subjective sleep quality across intervention visits. Boxplots show OSQ scores for Group 1 (blue; V1 baseline → V2 follow-up) and Group 2 (orange; V2 baseline → V3 follow-up). Group 2 showed a significant decrease in OSQ scores (p = 0.03), indicating improved subjective sleep quality, while Group 1 showed no significant change (p = 0.45). Lower scores reflect better perceived sleep quality.

### Physiological Data between Groups

No statistically significant differences in the analyzed sleep-related physiological variables were detected between Groups 1 and 2 during the digital monitoring phase (all p > 0.05); data were therefore pooled for subsequent exploratory analyses.

**Table 1.** Comparison of sleep-related physiological variables between Group 1 and Group 2 during the digital monitoring phase. Median values of sleep parameters derived from wearable sensors are presented for both groups. No statistically significant differences were detected between groups (all p > 0.05).

|  | **Median Group 1** | **Median Group 2** | **p-value** |
|---|---|---|---|
| Global sleep score | 75 | 75.7 | 0.41 |
| Respiratory rate (BPM) | 14.3 | 15.1 | 0.93 |
| RMSSD (ms) | 24 | 35 | 0.32 |
| NREM heart rate (bpm) | 59 | 58.5 | 0.13 |
| Entropy HRV (complexity of inter-beat intervals) | 2.60 | 2.70 | 0.52 |
| Total sleep duration (minutes) | 340 | 338 | 0.30 |
| Deep sleep duration (minutes) | 63 | 61 | 0.60 |
| REM sleep duration (minutes) | 80 | 75 | 0.30 |

### Physiological Data Clustering

Continuous physiological signals recorded during the digital monitoring phase, including nightly skin temperature and respiratory rate from the Fitbit Sense, were clustered to identify distinct physiological sleep profiles across participants.

**Nightly skin temperature clustering** identified three thermal profiles (Temp_Cluster 0: n=26; Temp_Cluster 1: n=40; Temp_Cluster 2: n=27). Temp_Cluster 0 maintained lower temperatures around 33.0°C, Temp_Cluster 1 showed intermediate values near 33.7°C, and Temp_Cluster 2 remained higher around 34.2°C, indicating distinct thermoregulatory profiles among participants.

**Figure 3.**
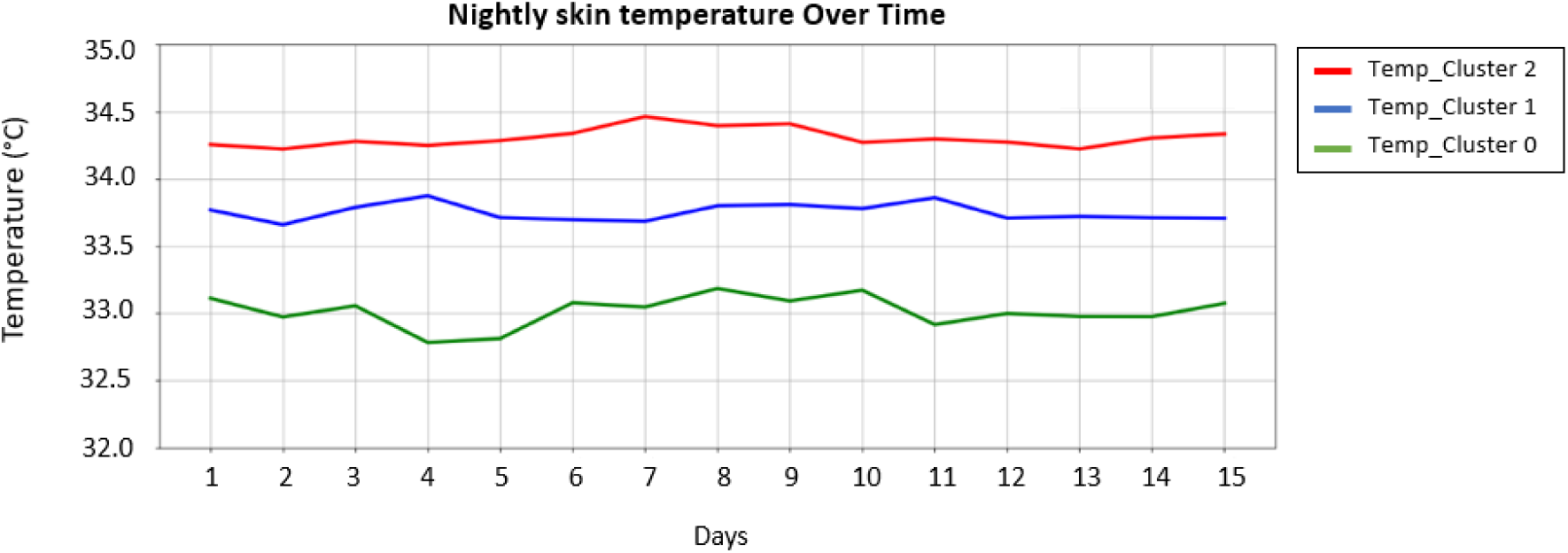
Average nightly skin temperature over the 15-day digital phase by thermoregulation cluster.

**Respiratory rate clustering** also revealed three distinct patterns (BR_Cluster 0: n=26; BR_Cluster 1: n=27; BR_Cluster 2: n=40). BR_Cluster 0 exhibited the lowest respiration rates (∼12-13 BPM), BR_Cluster 1 showed moderate rates (∼15-16 BPM), and BR_Cluster 2 presented higher and more stable rates (∼18-19 BPM). These patterns remained consistent across nights, reflecting individualized physiological sleep profiles./

**Figure 4.**
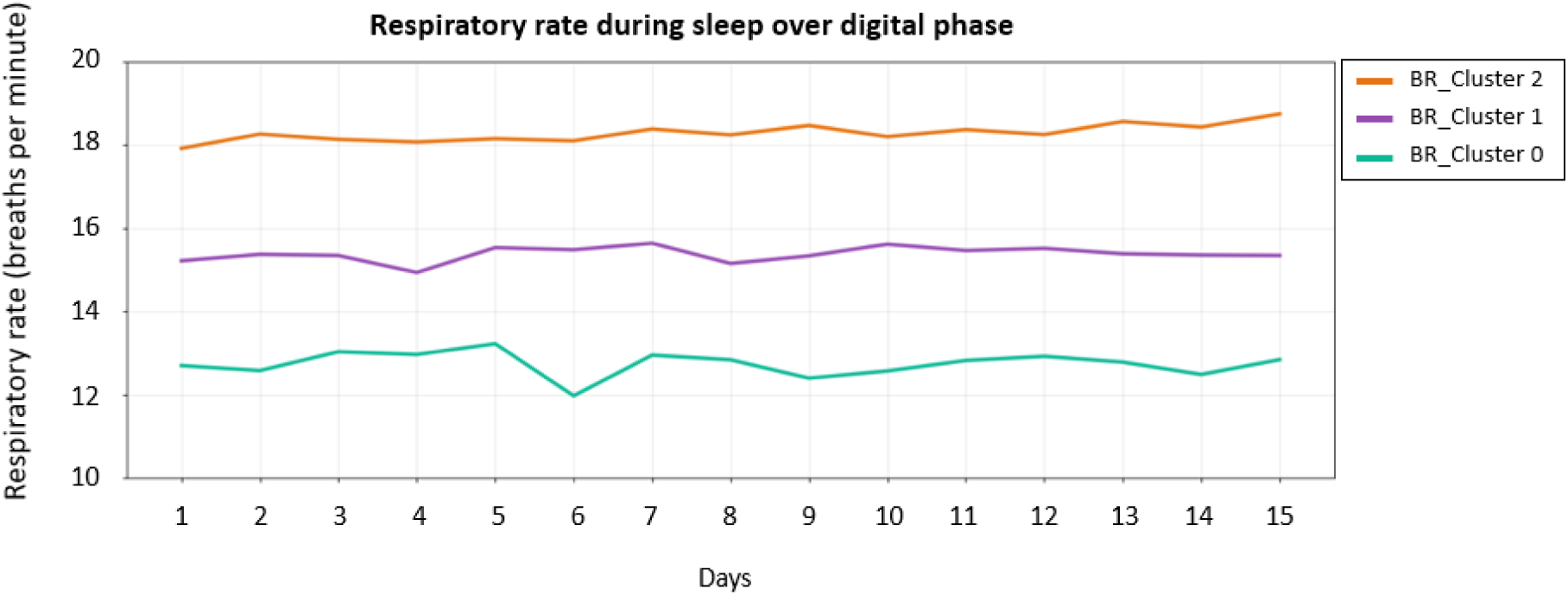
Average nightly respiratory rate over the 15-day digital phase by respiratory cluster.

### Nutritional Clusters

Nutritional data comprised 195 three-day dietary records from 92 participants. Participant-level nutritional features were subsequently analyzed using PCA and classified into three clusters:

- **Nut_Cluster 0** (n = 33): Low intake of most nutrients (deficiency profile).
- **Nut_Cluster 1** (n = 21): High nutrient density (elevated micro-and macronutrient intake).
- **Nut_Cluster 2** (n = 38): Balanced consumption (intermediate group).

Differences between clusters were particularly marked for fiber, potassium, folate, and vitamins.

**Figure 5.**
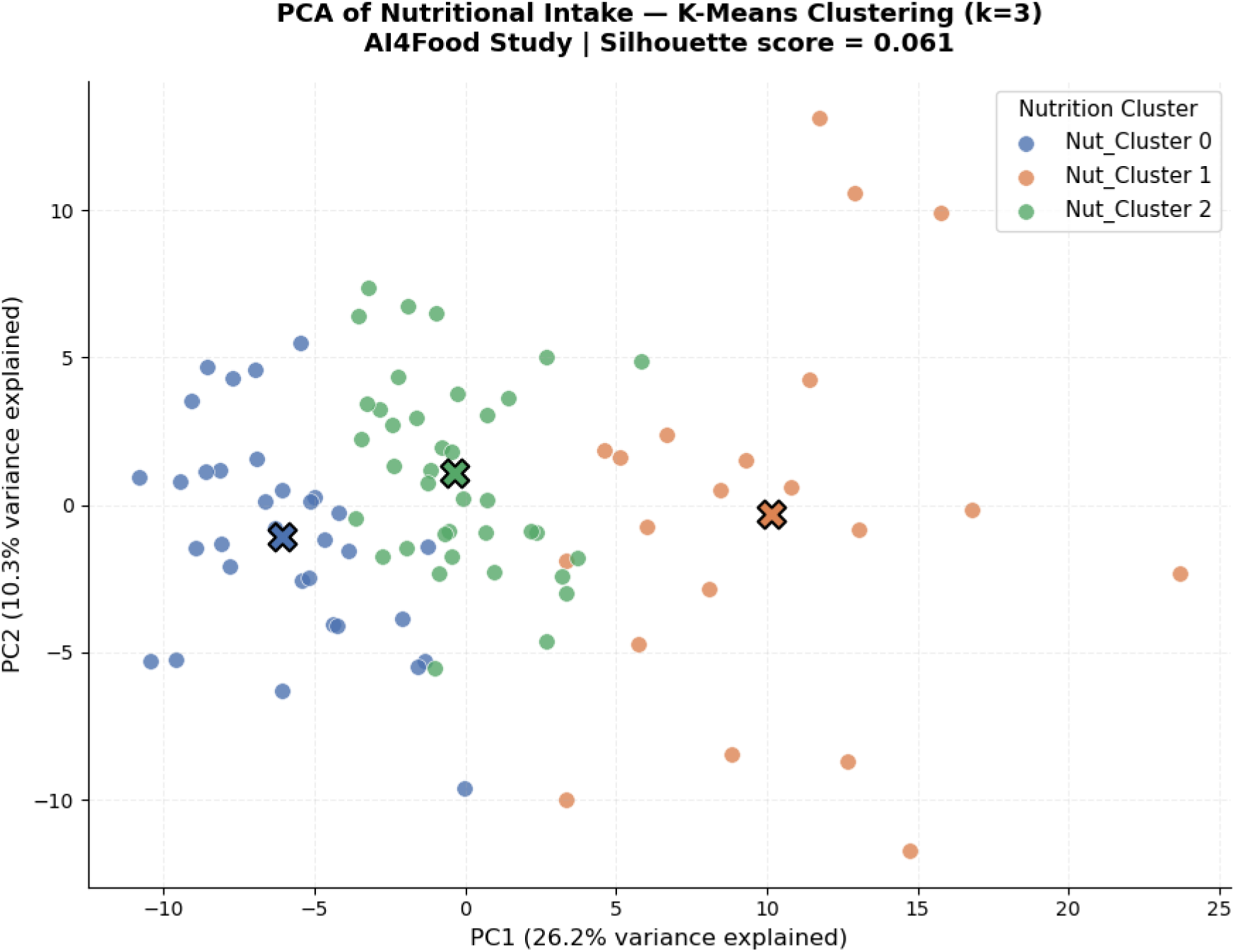
PCA of nutritional intake across participants, with k-means clustering. PC1 and PC2 together explain 36.5% of total variance (26.2% and 10.3%, respectively); cluster centroids are marked with X. Cluster separation was weak (silhouette score = 0.061), consistent with the overlap visible between clusters in PCA space.

**Figure 6.**
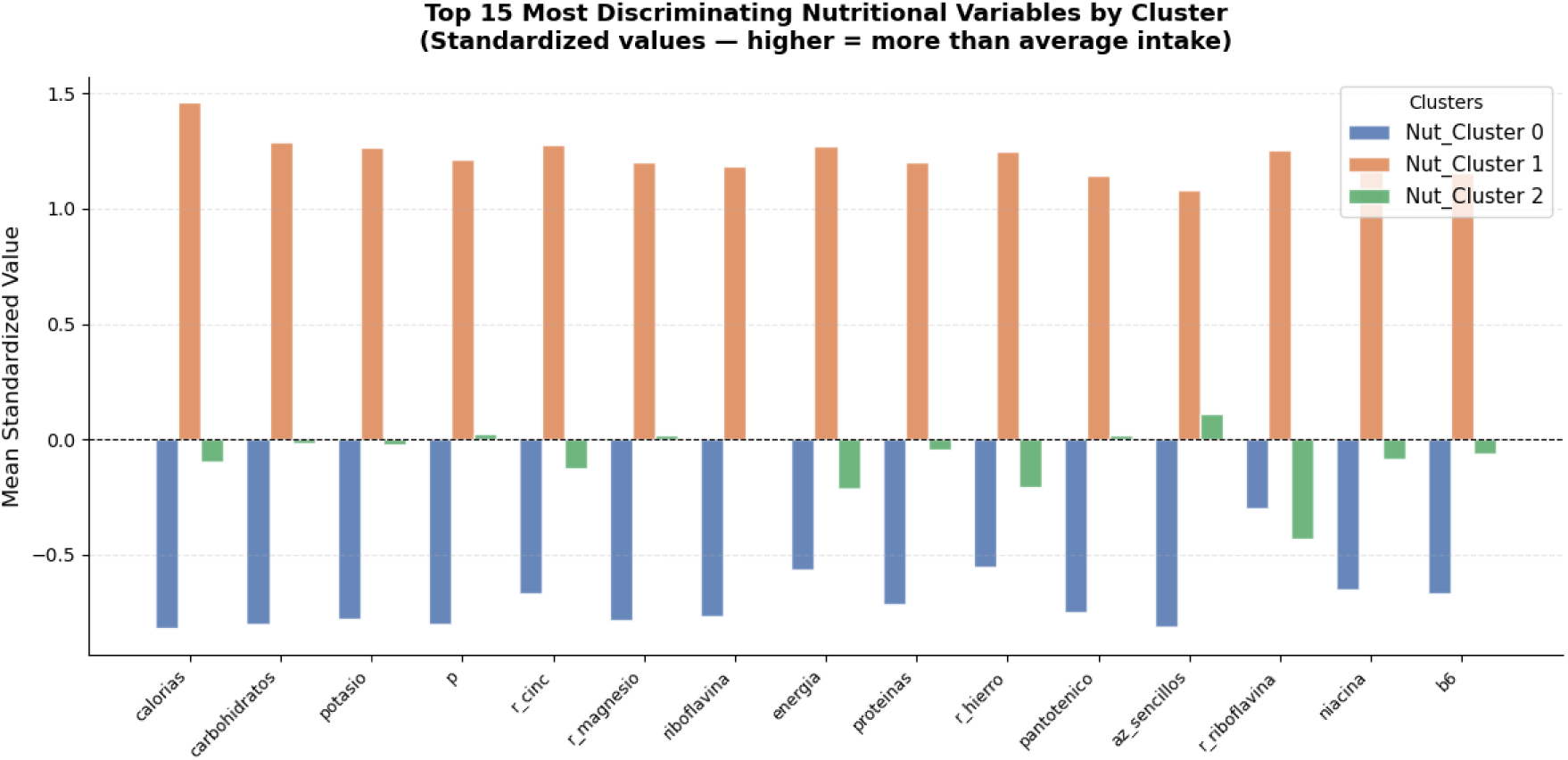
Top 15 most discriminating nutritional variables by cluster, expressed as mean standardized deviation (Z-score) from the global mean. Nut**_**Cluster 0 shows consistently lower-than-average intake across nearly all nutrients, while Nut_Cluster 1 shows consistently higher-than-average intake across the same variables. Nut_Cluster 2 remains close to the global average overall, with its largest deviations being lower r_riboflavina (≈−0.4) and lower iron and energy intake (r_hierro, energia; ≈−0.2 each), alongside a minor positive deviation in simple sugars (az_sencillos, ≈+0.1).

### Skin Temperature Dynamics

Nightly skin temperature differences (Δ°C over 24 h, relative to a session baseline) revealed two clear profiles. Both profiles showed an increase in wrist temperature during the nocturnal sleep window, consistent with established relationships between peripheral temperature rhythms, thermoregulation, and sleep (Kräuchi, 2007; Sarabia et al., 2008; Okamoto-Mizuno & Mizuno, 2012), followed by a decline after waking:

**Low temperature:** Strong nocturnal-diurnal amplitude (≈2.5°C drop after waking), reflecting a greater nocturnal–diurnal wrist-temperature amplitude.

**High temperature:** Minimal amplitude (mean ≈ −0.5 °C across the 24-h cycle, with an instantaneous nocturnal minimum reaching approximately −1.5 °C), reflecting a lower-amplitude wrist-temperature profile over the observed 24-hour period.

This separation was consistent across participants and days, as visualized by daily cluster heatmaps.

**Figure 7.**
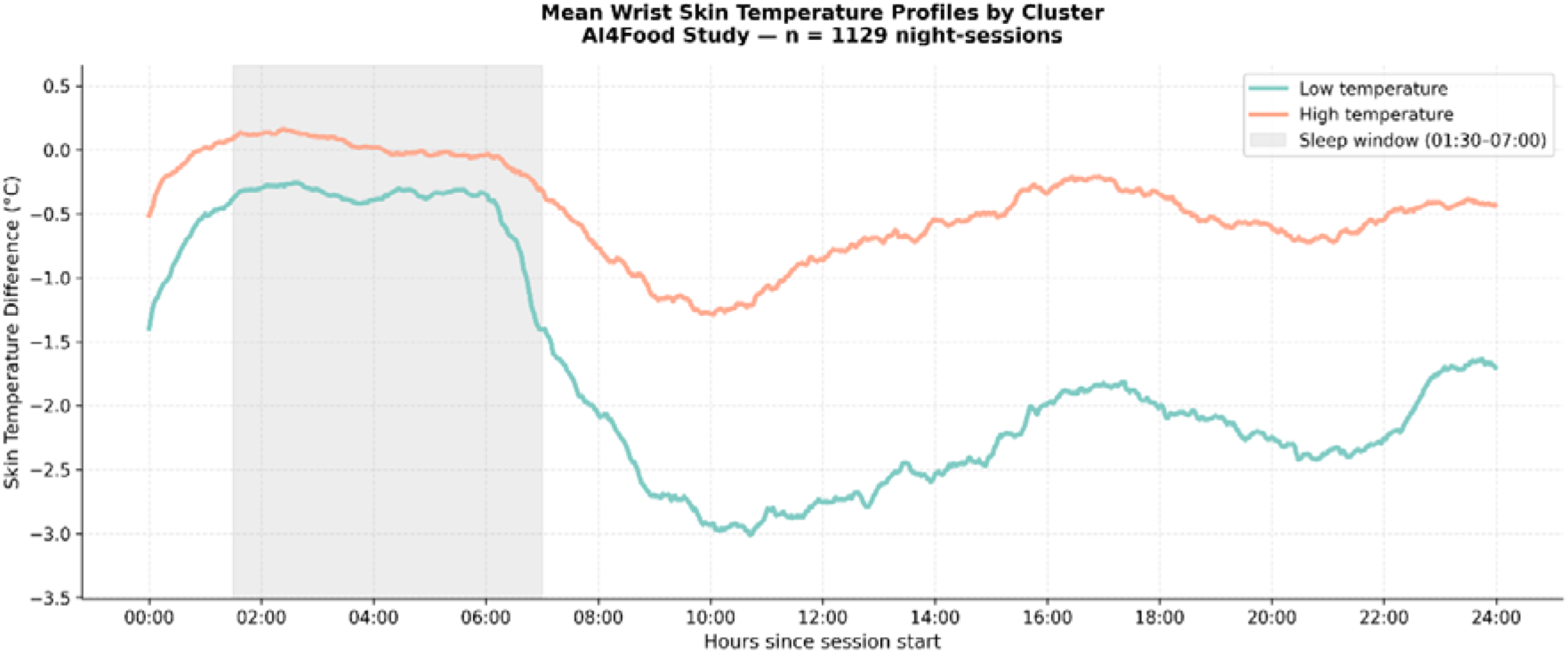
Wrist temperature difference aligned with the sleep period. Average wrist temperature variation (°C) over 24 hours for two temperature profiles. Both clusters showed an increase in wrist temperature during the shaded nocturnal sleep window (01:30–07:00), followed by a decline after waking. This decline was more pronounced in the Low temperature cluster (≈−2.5°C, 07:00–10:00), indicating a greater nocturnal–diurnal wrist-temperature amplitude.

**Figure 8.**
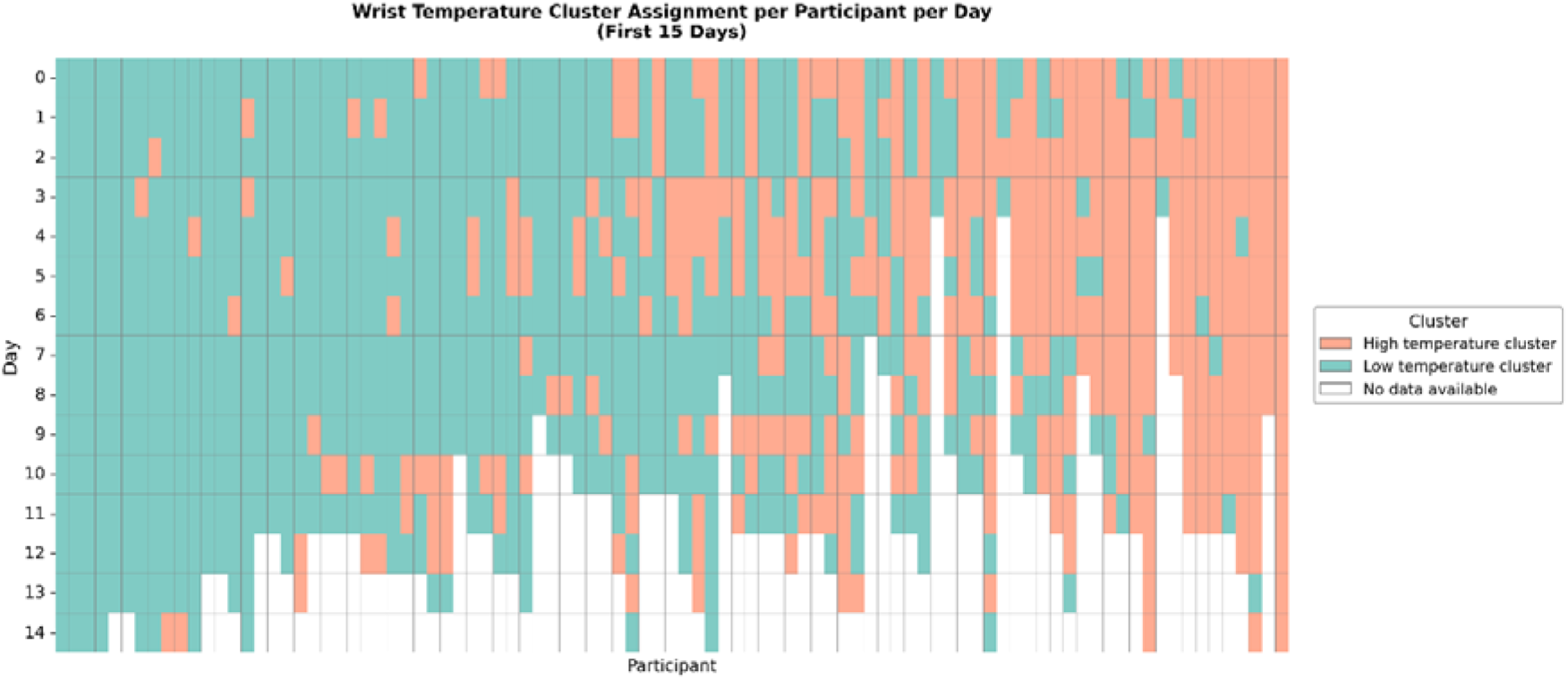
Participant cluster heatmap based on wrist temperature difference. Teal represents **Low temperature cluster** (higher amplitude), orange **High temperature cluster** (lower amplitude), and white indicates missing or excluded data. The heatmap shows daily cluster assignments for each participant over 15 days.

### Glucose Profiles

Continuous glucose monitoring identified two distinct 24-hour glycemic patterns:

**Low glucose:** Stable glucose rhythm with nocturnal minimum around 85 mg/dL.

**High glucose:** Higher baseline glucose with two daily peaks, a smaller rise in the morning and a more pronounced peak in the evening, resulting in a higher and more variable daily glycemic profile.

Individual heatmaps confirmed intra-participant stability of these clusters over time.

**Figure 9.**
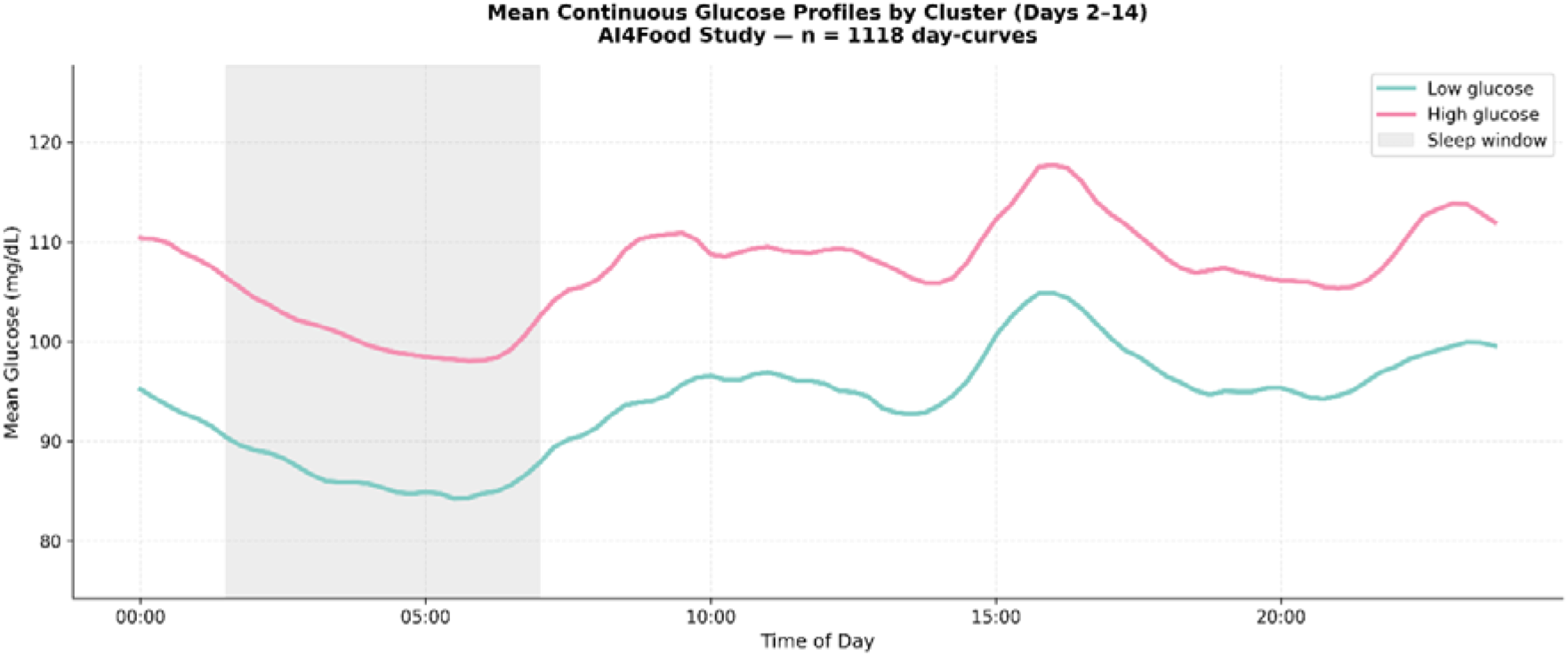
Glycemic level by cluster means over 24 hours. Average blood glucose levels (mg/dL) for two glycemic clusters across a 24-hour cycle. Low glucose cluster (teal) shows lower and more stable glucose values (∼85–105 mg/dL), while High glucose cluster (pink) maintains higher levels (∼95–115 mg/dL) with two daily peaks, a smaller morning rise and a more pronounced evening peak. The shaded area represents the sleep period.

**Figure 10.**
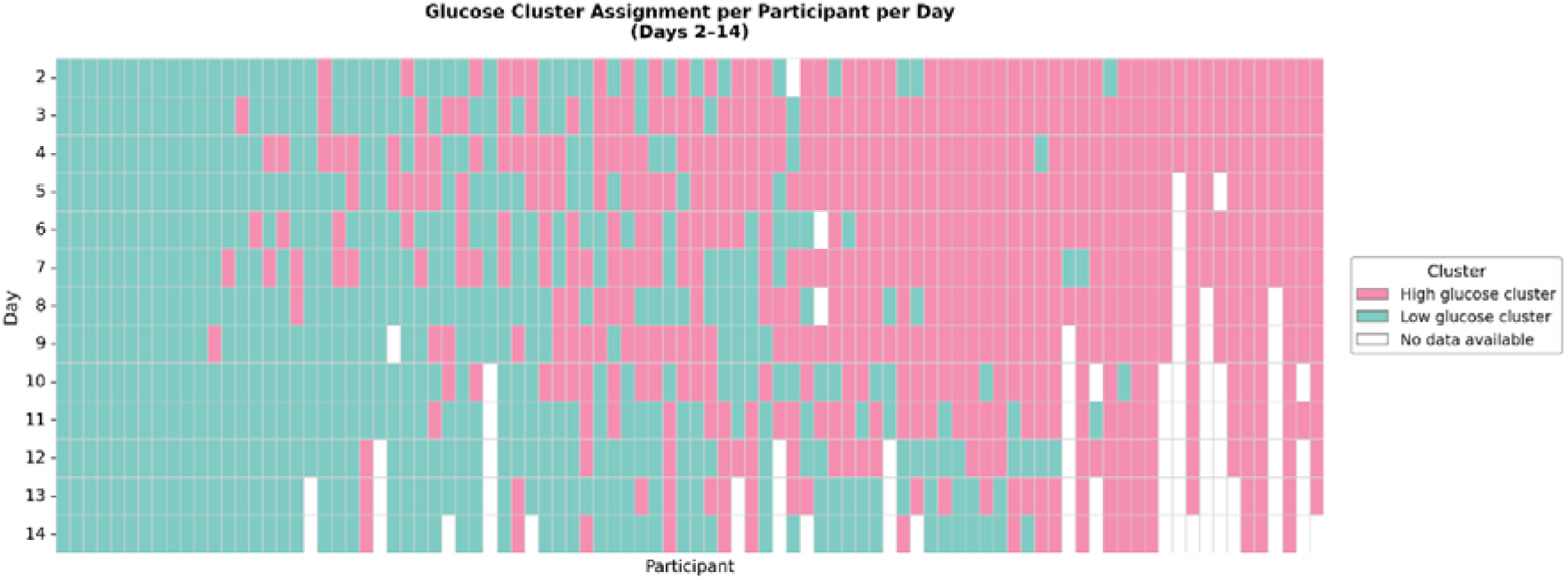
Heatmap representation of blood glucose clusters. Each cell shows the daily cluster assignment for a participant during the 15-day digital phase; the heatmap is shown through Day 13, as most participants had complete glucose recordings only up to that point, with later days affected by missing sensor data. Teal represents Low glucose cluster, pink High glucose cluster (higher glucose variability), and white indicates missing data. The heatmap highlights interindividual differences and temporal stability of glycemic patterns.

### Inter-Cluster Associations

To investigate whether physiological sleep profiles and dietary patterns were associated with weight-loss outcomes, cluster memberships across modalities were compared using contingency matrices and Chi-square tests. This approach allowed us to identify which physiological or nutritional phenotypes were more likely to co-occur with a given weight-variation profile.

No significant association was detected between nutritional and weight clusters (p = 0.57). However, significant associations were observed between:

- **Respiratory-rate clusters and weight clusters** (p < 0.01): Higher nocturnal respiratory-rate profiles were associated with smaller weight reductions.
- **Temperature clusters and weight clusters** (p = 0.01): Higher nighttime skin-temperature profiles were associated with smaller weight reductions.

These results suggest that sleep-related physiological markers may be associated with weight-loss responsiveness.

**Figure 11.**
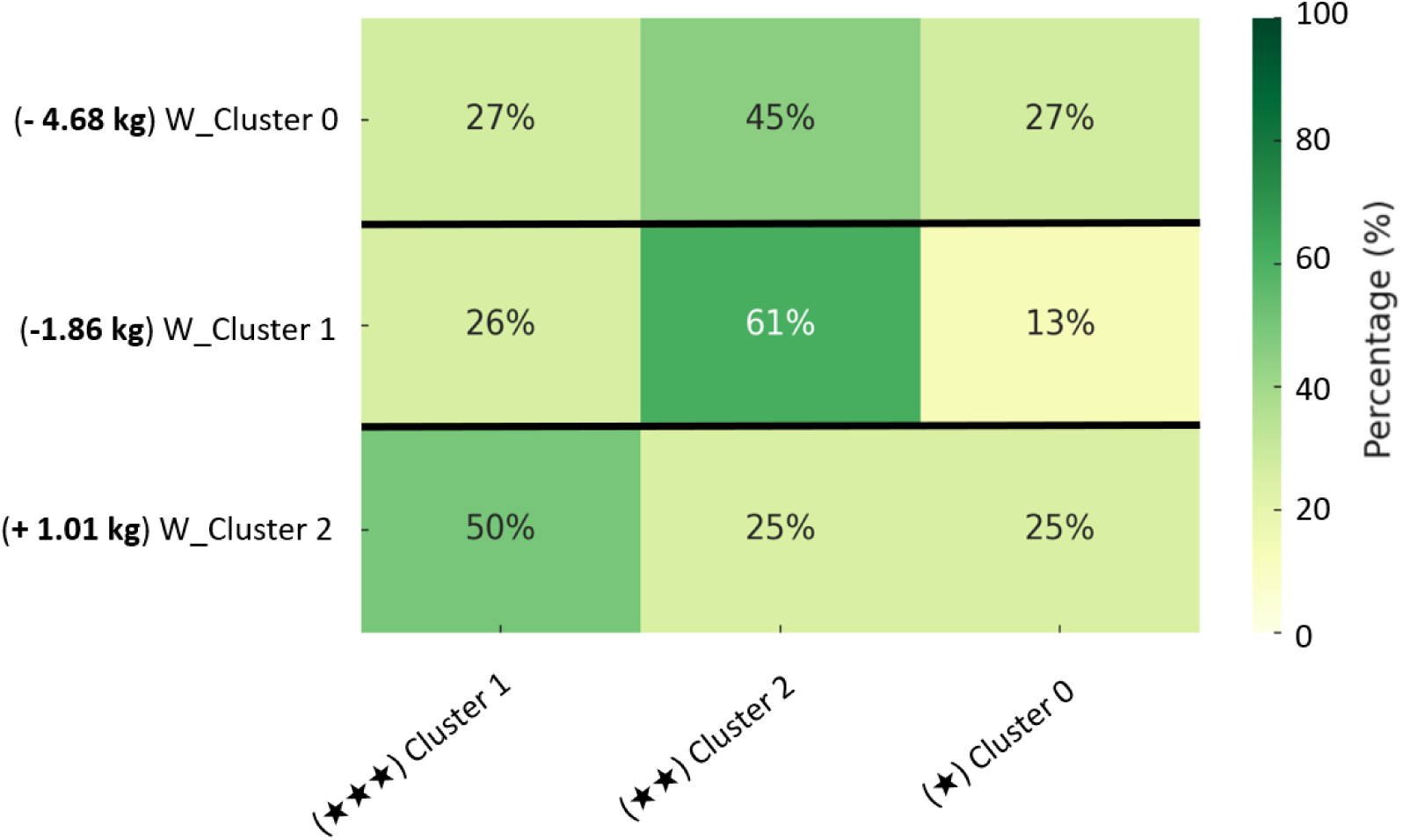
Relationship between weight and nutritional clusters (p = 0.57). The heatmap shows participant distribution (%) across nutritional intake levels: low (*), moderate (**), and high (***). No significant association was found between weight and nutritional clusters.

**Figure 12.**
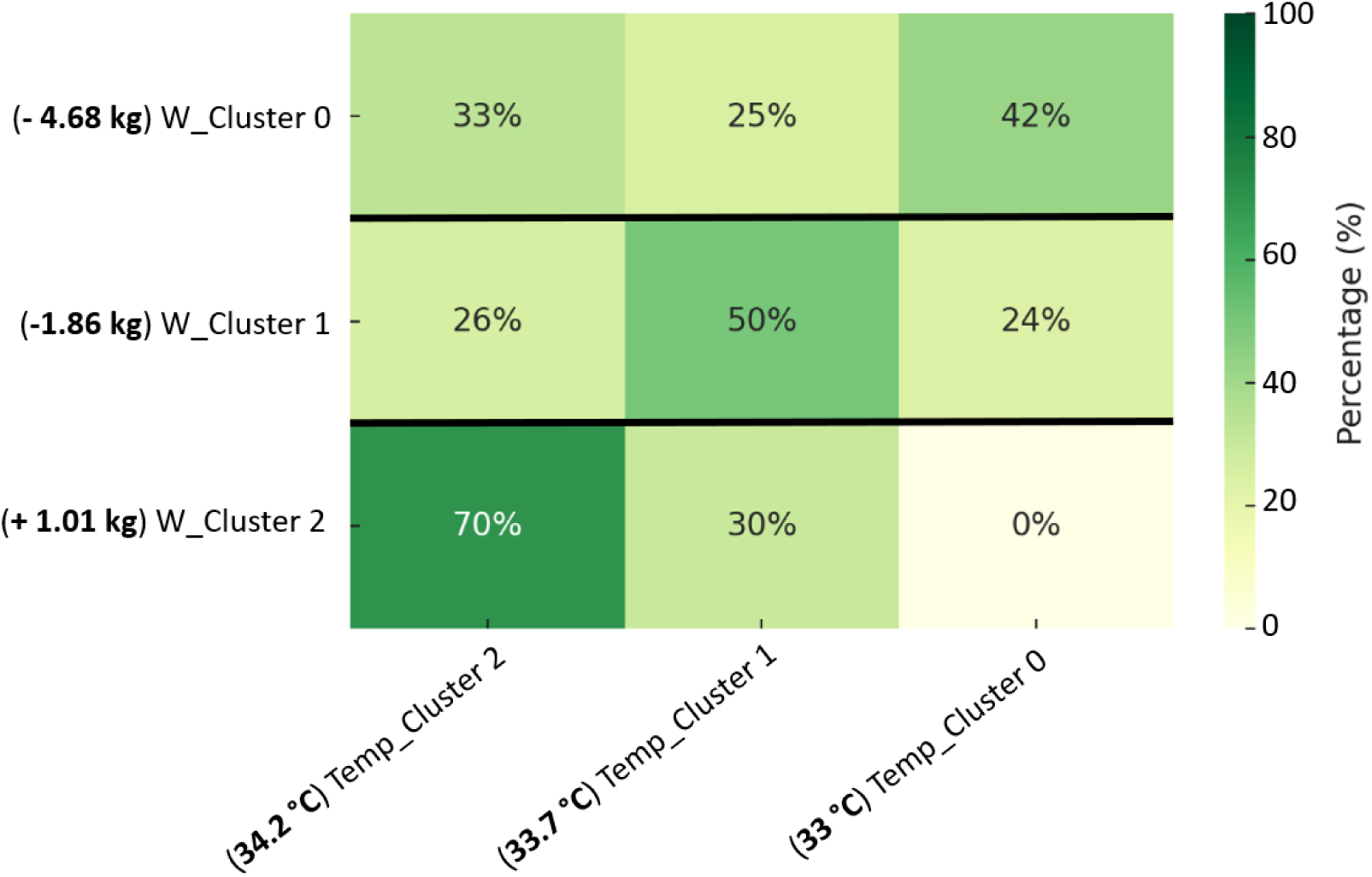
Relationship between weight and temperature clusters (p = 0.01). Participant distribution (%) across temperature clusters showed a significant association between temperature profile and weight-response cluster.

**Figure 13.**
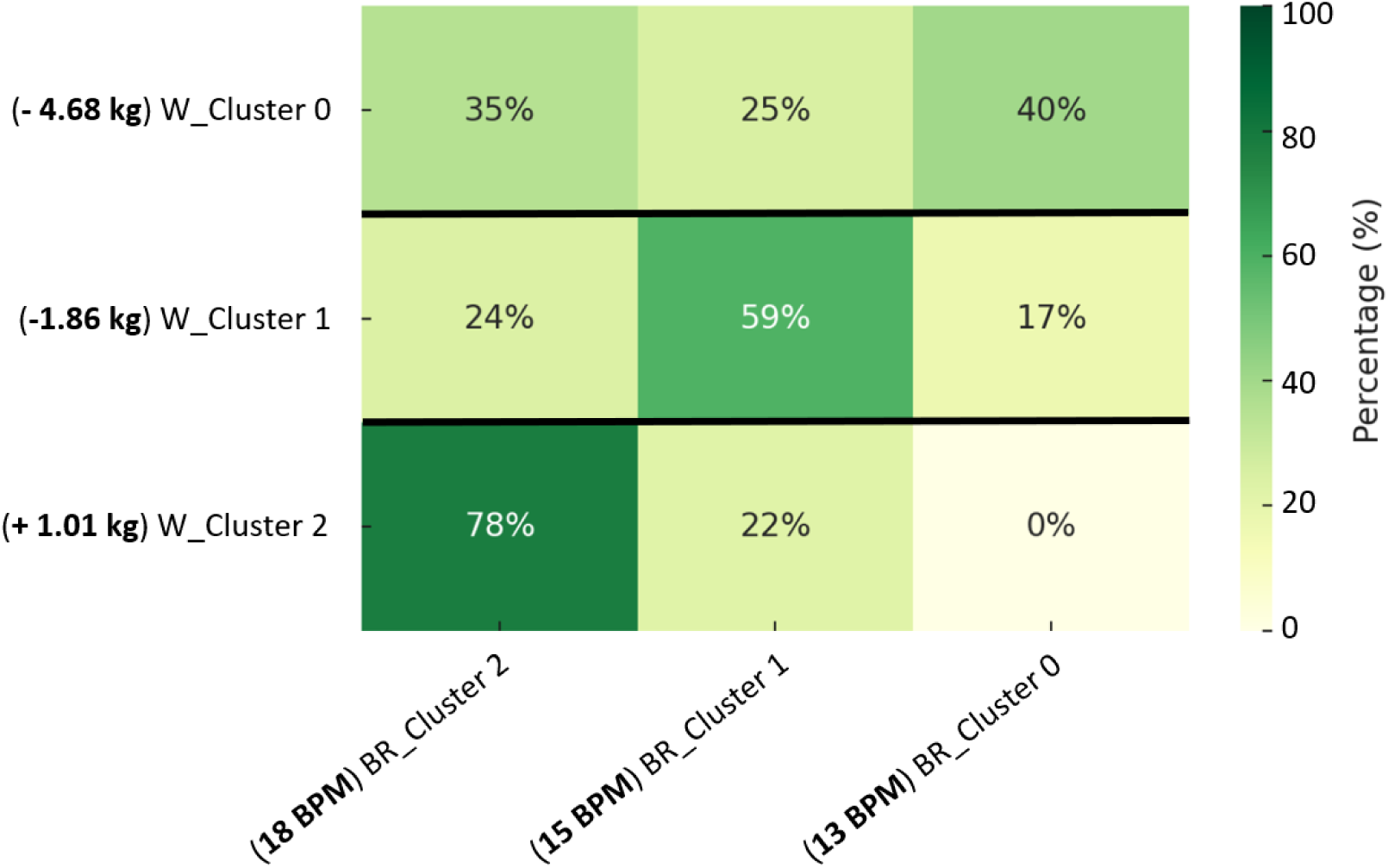
Relationship between weight and respiratory rate clusters (p *<* 0.01). Participant distribution (%) across respiratory-rate clusters showed a significant association between nocturnal respiratory profile and weight-response cluster.

### Predictive Modeling of Physiological Profiles

Participant-level five-fold cross-validation was used to evaluate the ability of sleep-related physiological variables to predict thermoregulation and glucose phenotypes. Random Forest and multilayer perceptron models were evaluated independently in each fold, with all observations from a given participant restricted to either the training or validation partition. Performance was summarized across folds using accuracy, precision, recall, F1-score, and ROC-AUC.

#### Model 1 – Prediction of Thermoregulation Phenotypes

Following consolidation to unique participant-day observations, the thermoregulation dataset comprised 261 observations from 55 participants. Fifteen non-temperature physiological predictors were included. Participant-level five-fold cross-validation was used to evaluate Random Forest and MLP classifiers. The Random Forest achieved a mean accuracy of 0.643 ± 0.057, mean F1-score of 0.453 ± 0.103, and mean ROC-AUC of 0.636 ± 0.034 across the five folds. The MLP achieved a mean accuracy of 0.518 ± 0.085, mean F1-score of 0.310 ± 0.205, and mean ROC-AUC of 0.558 ± 0.103.

**Table 2.** Pooled out-of-fold classification performance for thermoregulation phenotype prediction using Random Forest. Precision, recall, and F1-scores are shown for each phenotype, with predictions pooled across all held-out cross-validation folds.

|  | **Precision** | **Recall** | **Score F1** | **Support** |
|---|---|---|---|---|
| Low temperature | 0.60 | 0.849 | 0.739 | 152 |
| High temperature | 0.641 | 0.376 | 0.474 | 109 |
| **Accuracy** |  |  | 0.651 | 261 |

**Figure 14.**
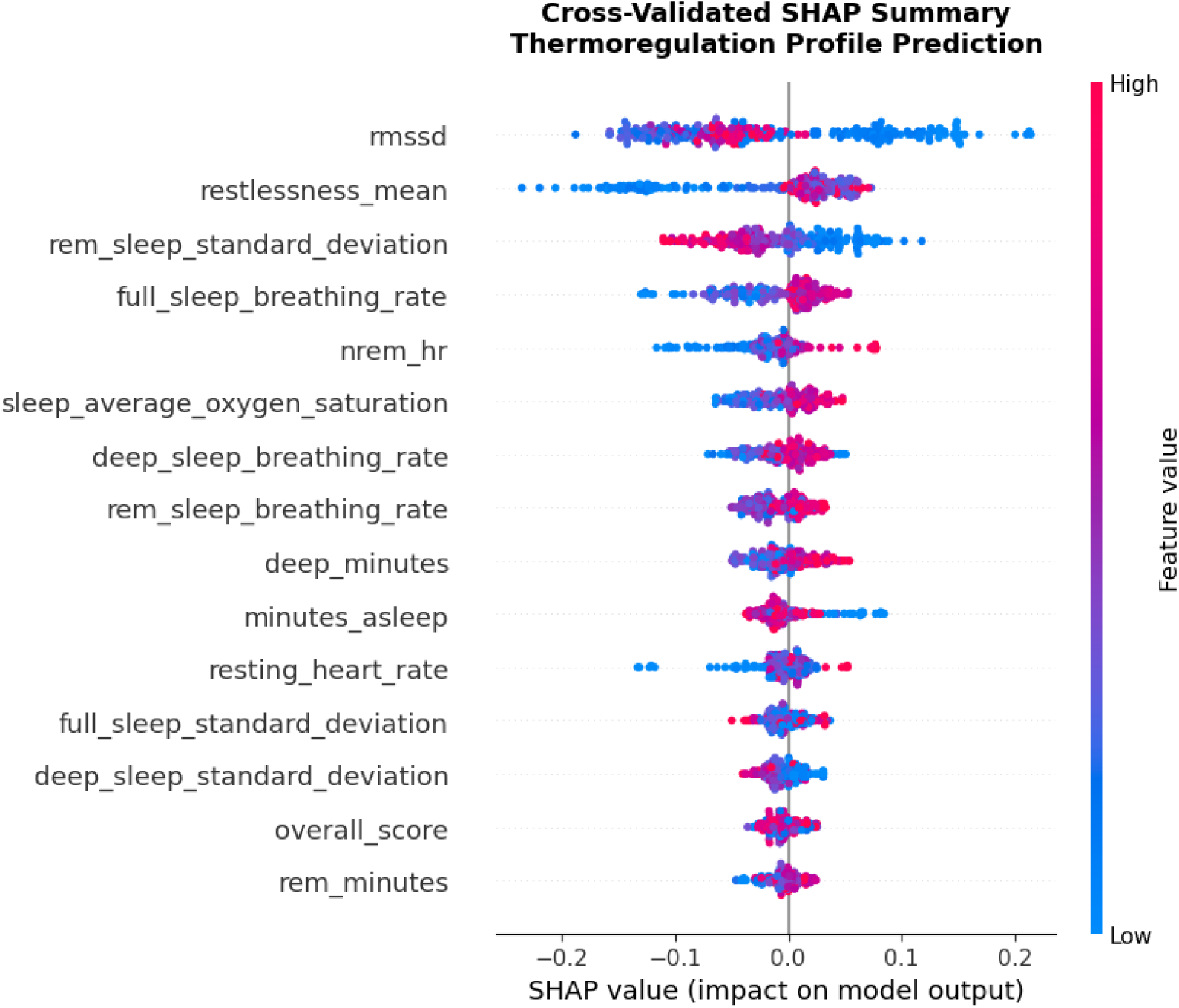
SHAP summary plot for thermoregulation prediction. RMSSD, restlessness, REM sleep respiratory-rate variability, full-sleep breathing rate, and NREM heart rate were the most influential predictors. Higher RMSSD tended to shift predictions toward the low-temperature profile, whereas greater restlessness and higher breathing rate tended to shift predictions toward the high-temperature profile.

#### Model 2 – Prediction of Glucose Phenotypes

Following consolidation to unique participant-day observations, the glucose prediction dataset comprised 144 observations from 44 participants. Seventeen raw sleep and physiological predictors were evaluated, while thermoregulation cluster membership was excluded from the predictor set. Under participant-level five-fold cross-validation, the Random Forest achieved a mean accuracy of 0.617 ± 0.113, mean F1-score of 0.492 ± 0.151, and mean ROC-AUC of 0.699 ± 0.170. The MLP achieved a mean accuracy of 0.438 ± 0.114, mean F1-score of 0.368 ± 0.225, and mean ROC-AUC of 0.448 ± 0.102.

**Table 3.** Pooled out-of-fold classification performance for glucose phenotype prediction using Random Forest. Precision, recall, and F1-scores are shown for each phenotype, with predictions pooled across all held-out cross-validation folds.

|  | **Precision** | **Recall** | **Score F1** | **Support** |
|---|---|---|---|---|
| High glucose | 0.551 | 0.429 | 0.482 | 63 |
| Low glucose | 0.621 | 0.728 | 0.670 | 81 |
| **Accuracy** |  |  | 0.597 | 144 |

**Figure 15.**
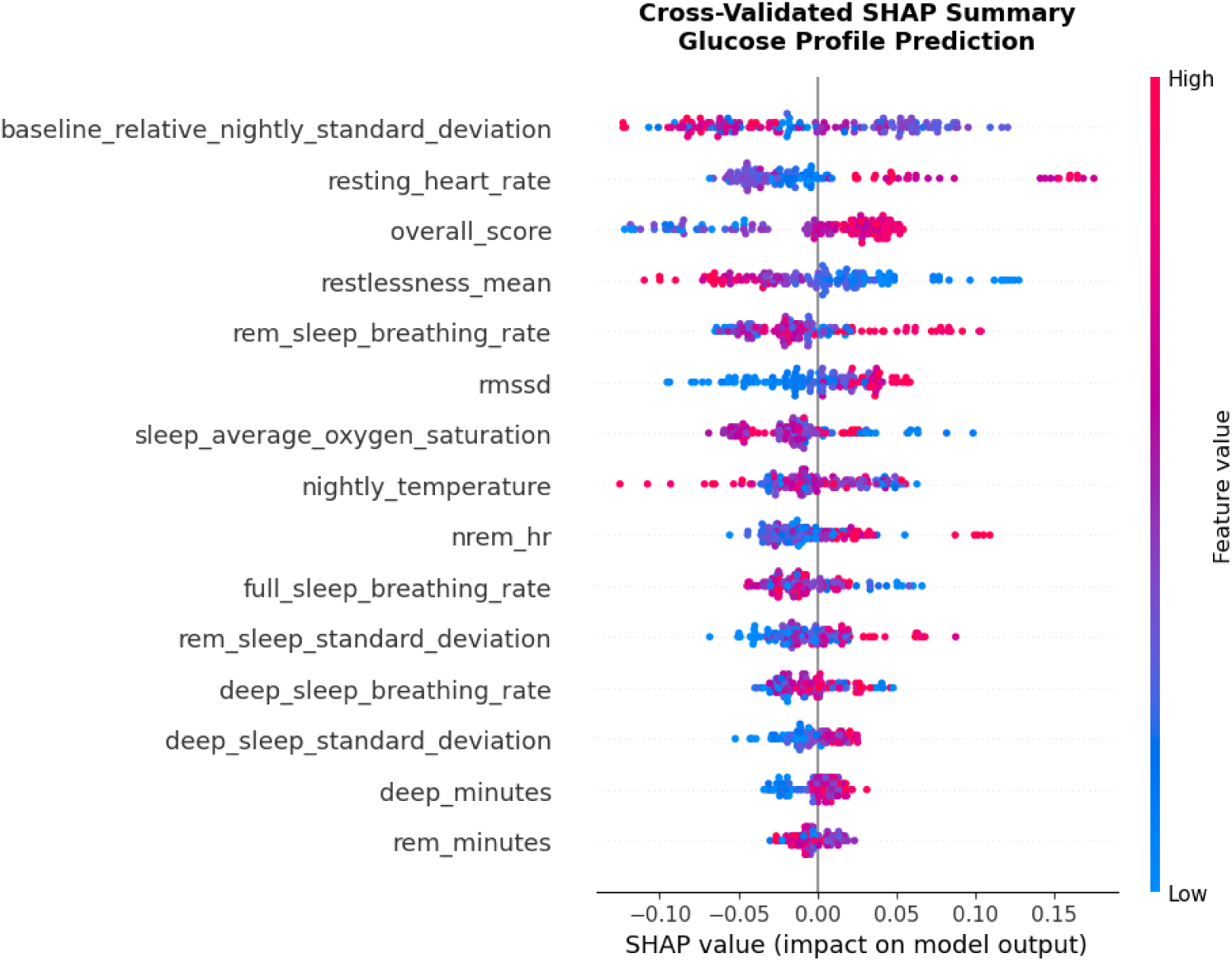
Cross-validated SHAP summary for glucose phenotype prediction. Baseline-relative nightly temperature variability, resting heart rate, overall sleep score, restlessness, and REM sleep breathing rate were among the most influential predictors. Each dot represents a held-out participant-day observation, with color indicating feature magnitude and SHAP values indicating the direction and magnitude of contribution to model predictions.

### Gut Microbiome Composition

Gut microbial composition was assessed at phylum, class, family, and genus levels between Visit 1 (V1) and Visit 3 (V3) across the three weight clusters. Communities were dominated by Bacteroidetes and Firmicutes at the phylum level, with Bacteroidaceae and Bacteroides remaining the most abundant family and genus across all groups. Minor shifts were observed, including a slight increase in Prevotellaceae in W_Cluster 0 and a trend toward increased Bacteroides with decreased Alistipes and Prevotella in W_Cluster 2, but none of these changes reached statistical significance.

The overall stability of gut microbiome composition across visits is consistent with previous studies showing that short-term dietary interventions are insufficient to induce substantial microbial shifts. The 30-day intervention period was likely too short to produce measurable or lasting changes in microbial community structure, and the observed trends should be interpreted with caution given the high interindividual variability inherent to microbiome data (Leeming et al., 2019).

**Figure 16.**
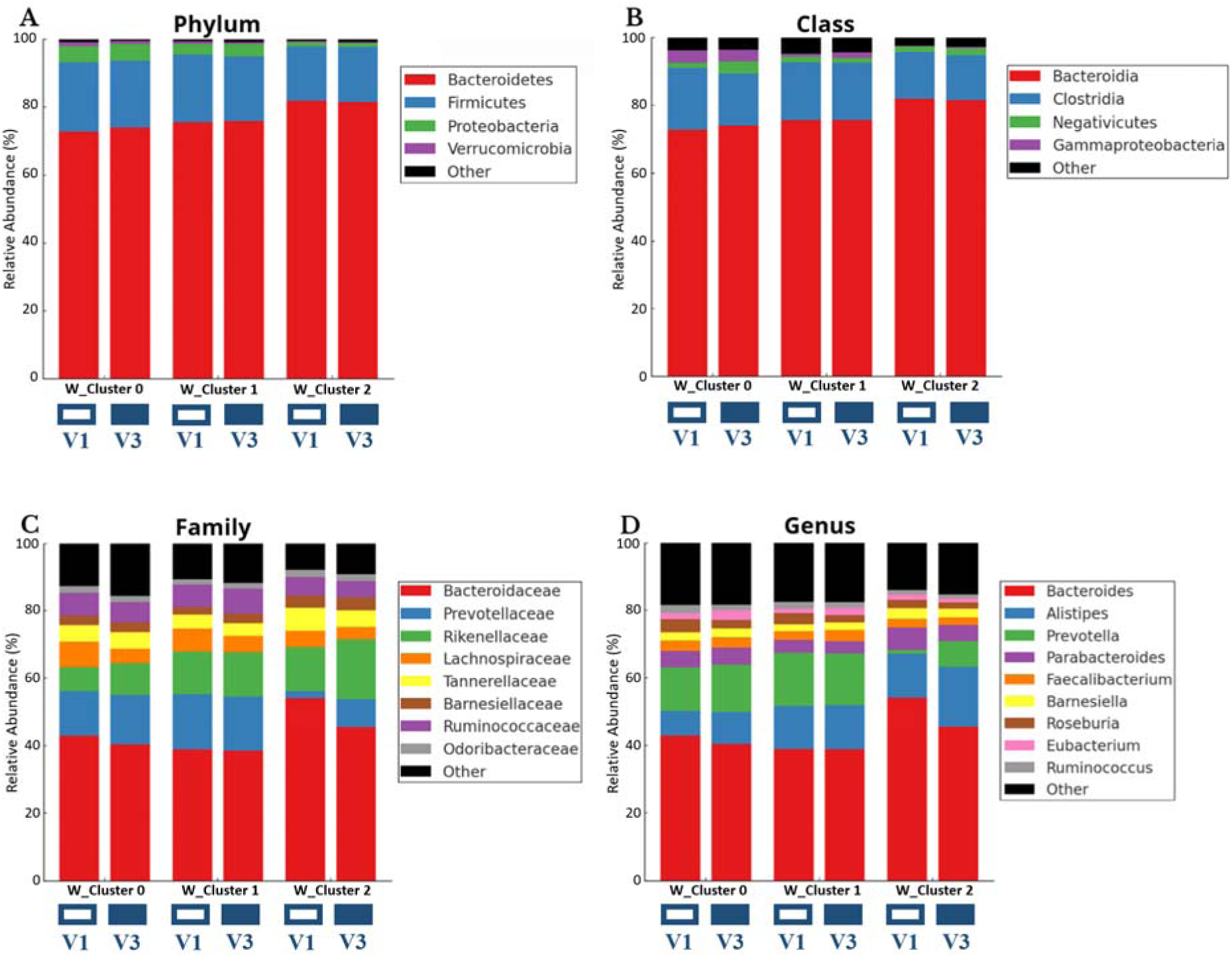
Gut microbiota relative abundance across weight clusters and visits at phylum, class, family, and genus levels. Panels (A–D) display bacterial community composition for W_Cluster 0 (−4.68 kg), W_Cluster 1 (−1.86 kg), and W_Cluster 2 (+1.01 kg) between Visit 1 (V1) and Visit 3 (V3).

## DISCUSSION

This study integrated wearable, nutritional, and biological data from a 30-day crossover intervention to investigate physiological heterogeneity in overweight and obese adults. The crossover design allowed direct within-participant comparison between monitoring modalities, and the overall intervention confirmed meaningful weight reduction across both groups, consistent with the outcomes reported in the parent AI4Food study (Lacruz-Pleguezuelos et al., 2025).

The observed trend toward greater weight loss during the digital monitoring phase aligns with the growing evidence that wearable-assisted health monitoring can support behavioral engagement in weight management (Dunn et al., 2018). However, since adherence and engagement were not directly measured, this interpretation should remain cautious. Subjective sleep quality improved significantly only in Group 2 during the traditional phase, highlighting that the effects of monitoring modality may differ across health outcomes and that digital monitoring does not uniformly improve all measured dimensions of health.

The clustering analyses revealed substantial interindividual variability in physiological and dietary profiles, which would not have been apparent from group-level summary statistics alone. The association between nocturnal respiratory-rate and wrist-temperature profiles with weight-variation clusters is consistent with the established role of thermoregulation and autonomic function in sleep regulation and metabolic health (Kräuchi, 2007; Okamoto-Mizuno & Mizuno, 2012; Sarabia et al., 2008). The link between sleep physiology and metabolic outcomes is further supported by evidence that sleep disturbances are associated with insulin resistance and increased risk of obesity (Reutrakul & Van Cauter, 2018). Nutritional cluster membership was not significantly associated with weight change, suggesting that dietary patterns alone, as captured by three-day records over a short intervention, may be insufficient to explain interindividual weight-loss variability.

Machine-learning analyses showed that sleep-derived physiological variables contain modest predictive information about thermoregulation and glucose phenotypes. Random Forest consistently outperformed the MLP, which is consistent with the known advantages of tree-based methods on structured tabular data with limited sample sizes (Breiman, 2001). The SHAP analysis identified autonomic and sleep-architecture variables as the most influential predictors, supporting the hypothesis that nocturnal physiology reflects broader metabolic differences. The link between wearable skin-temperature dynamics and physiologically meaningful interindividual differences has been similarly reported in recent work (Burks et al., 2026; Tai et al., 2023). For glucose phenotype prediction, the moderate Random Forest performance is in line with studies showing that wearable-derived signals can partially capture glycemic variability, though CGM-based features and larger cohorts improve discrimination substantially (Zeevi et al., 2015; Metwally et al., 2026).

Gut microbial composition remained stable across visits and was not significantly associated with weight-variation clusters. This finding is consistent with evidence that short dietary interventions are generally insufficient to produce sustained changes in gut microbiome structure, particularly given the high interindividual variability characteristic of microbiome data (Leeming et al., 2019).

Several limitations should be acknowledged. The effective sample size for predictive modeling was smaller than the full cohort, increasing uncertainty in performance estimates. Phenotype labels were generated before supervised cross-validation, which means the evaluation does not constitute a fully nested pipeline; future analyses should fit clustering independently within each training fold. The absence of a washout period, occasional missing wearable measurements, and the short intervention duration further constrain interpretation. P-values were not corrected for multiple comparisons, and all findings should be considered hypothesis-generating.

## Data Availability

The raw AI4FoodDB dataset is described in Romero-Tapiador et al. (2023) and is available at https://github.com/AI4Food/AI4FoodDB.

## ACKNOWLEDGEMENTS

This work was carried out within the **AI4Food-CM project** at the **IMDEA Nutrition Institute** (Madrid, Spain) under the supervision of **Dr. Enrique Carrillo de Santa Pau** and the **Computational Biology Group**, whose collaboration and expertise were invaluable. AI4Food-CM project is a collaborative work between the Computational Biology Group, the GENYAL Platform at IMDEA Nutrition, Madrid, Spain, and BiDA Lab group at Universidad Autónoma de Madrid, Spain.

## CODE AVAILABILITY

All code used for data processing, clustering, and predictive modeling in this study is publicly available at https://github.com/hicham-attioui/ai4food-metabolic-phenotyping.

## ABBREVIATIONS

AI4FoodDB: AI4Food Database
BMI: Body Mass Index
BPM: Breaths Per Minute
bpm: Beats Per Minute
CGM: Continuous Glucose Monitoring
CRP: C-Reactive Protein
ECG: Electrocardiogram
HbA1c: Glycated Hemoglobin
HRV: Heart Rate Variability
ML: Machine Learning
MLP: Multilayer Perceptron
ms: Milliseconds
NFC: Near Field Communication
NREM: Non-Rapid Eye Movement
OSQ: Oviedo Sleep Questionnaire
PCA: Principal Component Analysis
REM: Rapid Eye Movement
RMSSD: Root Mean Square of Successive Differences
ROC-AUC: Receiver Operating Characteristic - Area Under the Curve
SHAP: SHapley Additive exPlanations
SpO2: Oxygen Saturation
XAI: Explainable Artificial Intelligence

